# Determinants of normal haemoglobin concentration among children in Ghana: a positive deviance analysis of nationally representative cross-sectional survey data

**DOI:** 10.1101/19010769

**Authors:** Dickson A. Amugsi

## Abstract

Anaemia among children under 5, is a public health problem of serious concern. In Ghana, 8 out of every 10 children are anaemic. This study employed a novel approach to investigate the determinants of normal haemoglobin (Hb) concentration among children aged 6 to 59 months, using data from the Ghana Demographic and Health Surveys. The results showed that a year change in maternal education was positively associated with normal Hb concentration. Children of non-anaemic mothers were 1.67 (CI=1.32, 2.10; P<.001) times more likely to have normal Hb concentration relative to children of anaemic mothers. Compared to mothers who had less than 4 antenatal care (ANC) visits, mothers who had at least 4 ANC visits increased the odds of their children having a normal Hb concentration by 1.62 (CI=1.09, 2.40; P<.018). Children living in middle and rich households had respectively 1.48 (CI=1.06, 2.07; p<.021) and 1.59 (CI=1.08, 2.33; p<.018) increased odds of having a normal Hb concentration relative to those living in poor households. Maternal education, anaemia, ANC attendance, and household wealth index are strong determinants of normal Hb concentration among children in Ghana. Strategies aimed at addressing childhood anaemia should take into account maternal anaemia, education, poverty and ANC attendance.

## Introduction

Anaemia, defined as blood haemoglobin concentration lower than normal (1, 2) is a pervasive problem among children under five years of age. It reflects a state in which the number of red blood cells is low, or their ability to carry oxygen is poor (3). If anaemia is left untreated, it can have significant negative consequences on child health, some of which can be long lasting (4) and potentially derail the life-course wellbeing into adulthood. These adverse health consequences include but are not limited to poor cognitive development, difficulty with concentration, lethargy, increased mortality, susceptibility to infection and loss of economic productivity in adulthood (5, 6). Given the consequences, there is an urgent need for effective and efficient public health interventions to address anaemia in children. To be able to design effective interventions, it is significant to have clear understanding of the factors that have the potential to prevent children from developing anaemia. The present study is set out to illuminate these factors by focusing on children who have normal Hb concentration (i.e. children who do not have any form of anaemia). The study will provide a robust understanding of what promote normal Hb concentration, rather than the risk factors of low Hb concentration (anaemia). Indeed, knowledge about the determining factors of normal Hb concentration in children, which is currently lacking in sub-Saharan Africa (SSA), is essential for developing effective anaemia prevention programmes.

Data from the World Health Organisation (WHO) show that anaemia is one of the ten most serious health problems in the world (1). Globally, it is estimated that about 273.2 million children aged from 6 to 59□months suffer from anaemia, with an overall prevalence of 43% (3). The problem is particularly serious in Africa, where approximately 60% of preschool children are anaemic (7). The prevalence in the SSA region ranges from 42% in Swaziland to 91% in Burkina Faso (8). In Ghana, the focus of the present study, the overall prevalence of anaemia in children aged 6 to 59 months stands at 78.4% (9, 10), indicating that less than a third (21.6%) of children in Ghana have normal Hb concentration or are non-anaemic. The question is, what makes the 21.6% (defined in this study as *positive deviants*) to have normal Hb concentration, although they live in the same country as the rest of the children who are anaemic? What are the possible factors at the individual, household and community levels that influence normal Hb concentration for these children? Understanding this will help design effective programmes to help prevent pre-school children in Ghana from developing anaemia.

However, to be able to address the questions outlined above, we needed an approach that will focus on the positive aspects of health and the associated determinants rather than negative health outcomes. Therefore, this study utilised *positive deviance* (PD), a resource oriented approach as the analytical strategy. The concept of PD is premised on the notion that in “*every community there are certain individuals or groups whose uncommon behaviours and strategies enable them to find better solutions to problems than their peers, while having access to the same resources and facing similar or worse challenges”(11, 12)*. The PD concept aims to study the behaviours and characteristics of those who achieve better results on a given health outcome than their peers who reside in the same community (13). It is a well-established concept and can be explored using a statistical approach, and often quantified as those who do not experience a negative outcome of interest compared to those around them in the same settings (13). The PD approach has previously been used to investigate new born care, child nutrition, safe sexual practices, malaria control, health service delivery and educational outcomes in many settings (14-22). Indeed, almost all of these PD studies were conducted outside SSA. In Ghana for example, the application of PD in investigating child health outcomes is relatively rare. This suggests the need for more research using the PD approach in Ghana.

The novelty of the PD approach is its focus on ‘positive’ aspects of an outcome instead of the ‘negative’, and can identify potential points of intervention. In the present study, *positive deviants* were children who lived in anaemia endemic Ghana and yet have normal Hb concentration (not anaemic) relative to their counterparts who live in the same country but are anaemic. The main objective of this study is to examine the determinants of normal Hb concentration among children living in Ghana. Identifying these determinants would provide the basis for designing programmes, targeting children at risk of low Hb concentration in Ghana. This resource focused approach moves away from the dominant risk model approach, where the focus is usually on children with poor health and the associated risk factors.

### Methodology

#### Design and data sources

Data on haemoglobin (Hb) concentration (anaemia) among children aged 6–59 months from the 2014 Ghana Demographic and Health Surveys (GDHS) (23) were used in the analysis. The DHS surveys are designed to be representative at the national, regional and rural-urban levels. The Ghana DHS employed a two-stage sampling design. The first stage involved selection of clusters from a master sampling frame constructed from the 2010 national population and housing census. The clusters are selected using systematic sampling with probability proportional to size. The second stage selection involves the systematic sampling of the households listed in each cluster, and households to be included in the survey are randomly selected from the list. All women aged 15-49 years and their children aged 0-59 months were eligible to participate in the surveys. The data on children were obtained through face-to-face interviews with their mothers. The detailed description of the GDHS sampling strategy can be found elsewhere (10, 24). Our study sample comprised children aged 6–59 months (*n* =2451), born to mothers aged 15–49 years.

### Ethical considerations

The GDHS protocol, including biomarker collection, was reviewed and approved by the Ghana Health Service Ethical Review Committee and the Institutional Review Board of ICF International, USA. Written informed consent was obtained from participants before the survey. For children aged 6-59 months, consent was obtained from their parents or the adult responsible for the children. The GDHS team made sure that the biomarker results were made available to study participants. For example, the Hb test results for all children tested in the household were entered into a malaria and anaemia brochure and given to the parent or responsible adult. Finally, an anaemia referral form was given to facilitate immediate treatment at a nearby health facility of children with an Hb level less than 7.0 g/dl (10). The DHS Program, USA, granted the author permission to use the data. The data are completely anonymous; therefore the author did not seek further ethical clearance.

### Outcome and explanatory measures

#### Outcome measure

The main outcome measure for this analysis was Hb concentration (anaemia). The GDHS collected blood specimens for anaemia testing from all children aged 6-59 months. Blood samples were drawn from a drop of blood taken from a finger prick (or a heel prick in the case of children age 6-11 months) and collected in a microcuvette. The Hb analysis was carried out on-site using a battery-operated portable HemoCue analyser. The Hb data were categorised into; mild anaemia (Hb concentration of 10.0-10.9 g/dL), moderate anaemia (Hb concentration of 7.0-9.9 g/dL) and severe anaemia (Hb concentration of less than 7.0 g/dL) (10). In this analysis, mild, moderate and severe anaemia were recoded into “anaemic or low Hb concentration” and given a value of “0” (referenced category), and children who did not suffer from any form of anaemia were classified as “non-anaemic or normal Hb concentration” and given a value of “1” (outcome of interest). To make the interpretation of the results easier, considering the approach used, Hb concentration was used in place of anaemia in the interpretation of the results. Since *positive deviance* (25) is the analytic lens, children with normal Hb concentration were classified as *positive deviants*.

#### Explanatory measure

The explanatory measures were categorized into four levels of determinants: *child* (sex, age, birth order, diarrhoea, taking iron pills, drugs for intestinal parasites), *maternal* (age, education, anaemia status, antenatal care), *households* (household size, number of children under 5, wealth index) and *community* (place of residence). Some of these variables were recoded: antenatal care (ANC) was recoded into 0-3 visits (reference category) and 4 or more visits, while household wealth index was recoded into poor (reference category), and middle and richer as categories of interest. The selection of the explanatory variables was guided by the UNICEF conceptual framework of child care (26). The selected variables were further subjected to bivariate analysis to establish their relationship with the outcome variable (*results not shown*). All statistically significant variables were included in the multivariable logistic regression analysis.

#### Analytical framework and methods

The analysis in this paper was framed using the extended UNICEF conceptual framework for childcare (26, 27). The framework posits that child survival, growth and development are influenced by a web of factors, with three underlying determinants being food security, healthcare and a healthy environment, and care for children and women (26). These underlying determinants are in turn influenced by basic determinants. These basic determinants may be described as “exogenous” determinants, which influence child health through their effect on the intervening proximate underlying determinants. Thus, the underlying determinants are endogenously determined by the exogenous determinants (28). In this analysis, we built two empirical regression models of the determinants of normal Hb concentration. In the first model (Model 1), we included maternal, household and community level determinants. In the second and final model (Model 2), we adjusted for child level factors. The statistical significant level was set at p<.05. Results were presented as odd ratios (ORs) with 95% confidence intervals (CI).

## Results

### Descriptive analysis of the characteristics of the sample

The results in Table 1 show that 32% of children in Ghana have normal Hb concentration, while 56% of their mothers were non-anaemic or had normal Hb concentration. Twenty three percent (23%) of the children were reportedly taking iron pills/sprinkles/syrup at the time of the survey. A little over a third (38%) of the children received drugs for intestinal parasites. The results also showed that 13% of the children had diarrhoea during the last 2 weeks preceding the survey. An ANC attendance of at least 4 visits was high among mothers in the sample (89%). A little over half (54%) of the study participants were living in rural areas. The mean maternal number of years of education was 5.2±4.9, while the average age was 31.0±6.8 years. The mean child age was 31.3±15.4 months. Household size and child birth order were 5.8±2.7 and 3.4±2.1 respectively (Table 2).

**Table 1:** Characteristics of socio-demographic factors (n=2451), categorical variables

| Variables | % |
| --- | --- |
| *Child anaemia* |  |
| No anaemia | 32.2 |
| Mild to severe anemia | 66.8 |
| *Child sex* |  |
| Male | 53.1 |
| Female | 46.9 |
| *Maternal anaemia* |  |
| Not anaemic | 56 |
| Anaemic | 44 |
| Taking iron pills/sprinkles/syrup (yes) | 23.4 |
| Drugs for intestinal parasites (yes) | 37.6 |
| Had diarrhoea recently (yes) | 12.5 |
| *Place of residence* |  |
| Urban | 46.4 |
| Rural | 53.6 |
| *Number of antenatal attendance* |  |
| 0-3 visits | 11.2 |
| 4+ visits | 88.8 |
| *Household wealth index* |  |
| Poor | 42.9 |
| Middle | 19.9 |
| Rich | 37.2 |

**Table 2:** Characteristics of socio-demographic factors (n=2451), continuous variables

| Variables | Mean | SD |
| --- | --- | --- |
| Maternal education in years | 5.24 | 4.91 |
| Maternal age in years | 30.98 | 6.84 |
| Age of child in Months | 31.26 | 15.44 |
| Number of household members | 5.81 | 2.722 |
| Number of children under 5 years | 1.77 | .874 |
| Birth order of child | 3.37 | 2.09 |

214 SD=Standard deviation

### Multivariable analysis of the protective factors for childhood anaemia

The results in Table 3 show that all the maternal and household level factors except household size were significantly associated with normal Hb concentration in the first model (Model 1) but these significant associations disappeared after child level determinants were included the final empirical model (Model 2). Only maternal education, anaemia, ANC attendance, household wealth index and child age maintain their statistically significant association with normal Hb concentration in the final model. A unit change in maternal years of education was associated with 1.05 (CI=1.02, 1.08; P<.001) increased odds of normal Hb concentration. Children of mothers who had normal Hb concentration (non-anaemic) were 1.67 (CI=1.32, 2.10; P<.001) times more likely to have normal Hb concentration relative to children of anaemic (low Hb concentration) mothers. Compared to mothers who had less than 4 antenatal care (ANC) visits, mothers who had at least 4 ANC visits increased the odds of their children having a normal Hb concentration by 1.62 (CI=1.09, 2.40; P<.018). Additionally, children living in middle and rich households had 1.48 (CI=1.06, 2.07; p<.021) and 1.59 (CI=1.08, 2.33; p<.018) increased odds of having a normal Hb concentration respectively, relative to those living in poor households. A unit change in child’s age was associated with 1.03 (CI=1.02, 1.04; P<.001) increased odds of having a normal Hb concentration or not being anaemic.

**Table 3:** Multivariable analysis of the determinants of normal Hb concentration among children, 6-59 months (n=2451)

| Variables | Model 1 |  |  |  | Model 2 |  |  |  |
| --- | --- | --- | --- | --- | --- | --- | --- | --- |
|  | OR | 95% CI for OR |  | p | OR | 95% CI for OR |  | p |
| **Maternal level variables** |  |  |  |  |  |  |  |  |
| Maternal education (in years) | 1.050 | 1.023 | 1.078 | .001 | 1.051 | 1.022 | 1.080 | .001 |
| Maternal age (in years) | 1.026 | 1.009 | 1.043 | .002 | 1.021 | .995 | 1.047 | .110 |
| *Maternal anaemia* |  |  |  |  |  |  |  |  |
| Anaemic (ref) | ref | ref | ref | ref | ref | ref | ref | ref |
| Not anaemic | 1.581 | 1.263 | 1.980 | .001 | 1.667 | 1.324 | 2.101 | .000 |
| *Antenatal attendance* |  |  |  |  |  |  |  |  |
| Number of visits, 0-3 (ref) | ref | ref | ref | ref | ref | ref | ref | ref |
| Number of visits, 4+ | 1.558 | 1.058 | 2.294 | .025 | 1.616 | 1.087 | 2.402 | .018 |
| **Household level variables** |  |  |  |  |  |  |  |  |
| Number of household members | .966 | .912 | 1.022 | .227 | .970 | .915 | 1.028 | .305 |
| Number of children under 5years | .747 | .622 | .898 | .002 | .908 | .752 | 1.096 | .316 |
| *Household wealth index* |  |  |  |  |  |  |  |  |
| Poor | ref | ref | ref | ref | ref | ref | ref | ref |
| Middle | 1.530 | 1.109 | 2.110 | .010 | 1.482 | 1.060 | 2.072 | .021 |
| Rich | 1.666 | 1.149 | 2.415 | .007 | 1.591 | 1.084 | 2.334 | .018 |
| **Community level variables** |  |  |  |  |  |  |  |  |
| *Place of residence* |  |  |  |  |  |  |  |  |
| Rural (ref) | ref | ref | ref | ref | ref | ref | ref | ref |
| Urban | .827 | .616 | 1.110 | .206 | .829 | .611 | 1.124 | .227 |
| **Child level variables** |  |  |  |  |  |  |  |  |
| Age of child (in months) | .... | .... | ..... | ..... | 1.031 | 1.022 | 1.040 | .000 |
| *Sex of child* |  |  |  |  |  |  |  |  |
| Female (ref) | .... | .... | ..... | ..... | ref | ref | ref | ref |
| Male | .... | .... | ..... | ..... | 1.191 | .953 | 1.489 | .124 |
| *Taking iron pills/syrup* |  |  |  |  |  |  |  |  |
| No(ref) |  |  |  |  | ref | ref | ref | ref |
| Yes | .... | .... | ..... | ..... | .929 | .713 | 1.210 | .584 |
| *Drugs intestinal parasites* |  |  |  |  |  |  |  |  |
| No (ref) |  |  |  |  | ref | ref | ref | ref |
| Yes | .... | .... | ..... | ..... | .995 | .778 | 1.273 | .968 |
| *Had diarrhoea recently* |  |  |  |  |  |  |  |  |
| No (ref) |  |  |  |  | ref | ref | ref | ref |
| Yes | .... | .... | ..... | ..... | 1.222 | .877 | 1.701 | .236 |
| Birth order of child | .... | .... | ..... | ..... | .937 | .853 | 1.030 | .179 |

OR= Odds ratio; CI= confidence interval; ref=reference category; Hb=Haemoglobin

## Discussion

This study examined the determinants of normal Hb concentration among children in Ghana, using nationally representative data. The analysis focused on children who are doing well (*positive deviants*) rather than the usual practice of focusing on at risk or children with poor health outcomes. The results showed that maternal education, anaemia, ANC attendance, household wealth index and child age are strong determinants of normal Hb concentration among children. Mothers who have more years of education are likely to have children with normal Hb concentration. This suggests that the more years of education a mother has the likelihood that their children will have normal Hb concentration. This could be attributed to positive influence of education on maternal caregiving practices. There is evidence that mothers with more years of education are likely to be good caregivers and sensitive and responsive to caregiving duties (29, 30). They also tend to have disposal income to be able to provide nutritious, including iron rich food for their children. The evidence to this effect can be found in a study by Choi and colleagues (31), where children of educated mothers were reportedly consuming more iron rich diet such as meat and poultry than did children of less educated mothers. Educated mothers are also more likely to utilize child health services, which can have a positive effect on their children health outcomes (32, 33). The findings from the present study are consistent with the literature. Several studies have demonstrated that children with more educated mothers are less likely to develop anaemia than those with less educated mothers (31, 34-36).

Maternal anaemia status was also illuminated as an important determinant of normal Hb concentration among preschool children. There is a strong observed relationship between maternal anaemia status and child Hb concentration. Mothers who are non-anaemic turn to have children who have normal Hb concentration. Thus, whether a child will have normal Hb concentration or not is conditional on the anaemia status of the mother. This may be the case because poor maternal health has the potential to compromise optimal child caring practices, which will in turn have negative impact on their children health and wellbeing (37, 38). Also, mothers who are non-anaemic are likely to have access to nutritious food. Since there is a strong correlation between maternal and child dietary intake (39, 40), the likelihood that the children will also consume nutritious food is high. The literature on the effects of maternal anaemia status on childhood anaemia is abound. Studies have shown that mothers Hb level had significant effect on their children Hb levels (36, 41, 42). Furthermore, in Mozambique, the odds of having non-anaemic child was higher in communities with low percentage of anaemic mothers (43). Thus, strategies for minimising childhood anaemia must address maternal anaemia.

This study reveals that mothers who were able to attend at least four ANC visits are likely to have children with normal Hb concentration. This suggests that higher ANC visits has the potential to promote normal Hb concentration among children living in highly endemic anaemia settings. Therefore, promoting ANC attendance among women of reproductive age will have the potential to protect their children from developing anaemia in the first 5 years of their life. The positive effect of ANC attendance on childhood anaemia status could be attributed to the health and nutrition education mothers received during ANC visits. This could have positive impact on their feeding and caring practices, and consequently better health outcomes for their children. Kuhnt and Vollmer (44) found in their study that having at least four ANC visits is associated with reduced odds of malnutrition among children. ANC attendance has also been found to associate directly with improved birth outcomes and longer-term reductions of child mortality and malnourishment (44). There is evidence that women who have higher ANC visits are also likely to patronise postnatal services (45), with the consequential positive effect on their children health outcomes. Our findings together with literature suggest that interventions to address childhood anaemia in Ghana should promote ANC use among women of reproductive age, so as to protect their children from developing anaemia.

Similarly, the analysis show that the level of household wealth is an important determinant of normal Hb concentration among children in Ghana. Children who live in middle and rich household are more likely to have a normal Hb concentration relative to those who live in poor households. The association is stronger in the rich households than the middle class households. The differential effect can be explained by the fact that children in rich households are likely have easy access to nutritious food and better caring practices, prerequisites for optimal child health. The effect of household wealth on normal Hb concentration could be a confirmation of the widely recognised health benefits of living in better-off households (42, 46-48). Conversely, Agho and colleagues (49) observed that children from the richest and middle-class households had a lower Hb concentration than those from the poorest households (49). This negative relationship notwithstanding, the evidence as highlighted above is that children in wealthy household tend to have higher Hb concentration or better health outcomes compared to those living in poor households.

The study is associated with some strengths and limitations. The use of large nationally representative sample makes the observed associations more robust, and enhanced the generalisability of the findings to all children 6-59 months in Ghana. Further, the outcome variable was objectively measured, reducing possible misclassification. The novelty of this study is its focus on non-anaemic (normal Hb concentration) children and the associated protective factors rather than risks factors for childhood anaemia. A limitation worth mentioning is the cross-sectional nature of the data, which does not lends itself to the establishments of causal relationship between the predictor and outcome variables. The conclusions in the paper are therefore interpreted as mere associations between the predictor variables and the outcome variable. Another limitation is that the use of PD is somewhat limited as we were not able to explore all the potential PD behaviours that may contribute to normal Hb concentration in Ghanaian children using quantitative data of this nature. Notwithstanding, PD is a well-established concept and hence makes it possible to explore the approach using statistical methods.

## Conclusions

The results of study showed that mothers who have higher years of education are likely to have children with normal Hb concentration. This suggests that the more years of education a mother may potentially results in their children having normal Hb concentration. There is a strong relationship between maternal anaemia status and child Hb concentrations. Mothers who are non-anaemic turn to have children who have normal Hb concentration. Children who live in middle and rich household are more likely to have a normal Hb concentration relative to those who live in poor households. Mothers who are able to attend at least four ANC visits are likely to have children with normal Hb concentration. Thus, higher ANC visits has the potential to protect children living in highly endemic anaemia settings from developing anaemia. Interventions aimed at addressing childhood anaemia in Ghana should take into account maternal anaemia, education, poverty and ANC attendance.

## Data availability statement

This study was a re-analysis of existing data that are publicly available from The DHS Program at http://dhsprogram.com/publications/publication-fr221-dhs-final-reports.cfm. Data are accessible free of charge upon a registration with the Demographic and Health Survey program (The DHS Program). The registration is done on the DHS website indicated above.

## Acknowledgements

**I** wish to express our profound gratitude to The DHS Program, USA for providing us access to the data. I also wish to acknowledge Ghana Statistics Service and Ghana Health Service which played critical roles in the data collection process. My gratitude to Dr. Razak Gyasi for taking the pain to review the final manuscript.

## Authors’ Contribution

D.A.A., conceived and designed the study, performed the data analysis, interpreted the results and wrote the manuscript. The author takes responsibility of any issues that might arise from the publication of this manuscript.

